# Multiple vaccine comparison in the same adults from the VITAL study reveals vaccine-specific and age-related humoral response patterns

**DOI:** 10.1101/2024.01.22.24301601

**Authors:** Marieke van der Heiden, Sudarshan Shetty, Elske Bijvank, Lisa Beckers, Alper Cevirgel, Yannick van Sleen, Irina Tcherniaeva, Thierry Ollinger, Wivine Burny, Rob S van Binnendijk, Marianne A van Houten, Anne-Marie Buisman, Nynke Y. Rots, Josine van Beek, Debbie van Baarle

## Abstract

Vaccine responsiveness is often reduced in older adults. Yet, our lack of understanding of low vaccine responsiveness hampers the development of effective vaccination strategies to reduce the impact of infectious diseases in the ageing population.

Young-adult, middle-aged and older-adult participants of the VITAL clinical trials (n=315, age range: 28-98y), were consecutively vaccinated with a booster quadrivalent influenza (QIV) vaccine, a primary 13-valent pneumococcal-conjugate (PCV13) vaccine, and a primary series of SARS-CoV2 mRNA-1273 vaccines within the timeframe of 2 years. This unique setup allowed investigation of humoral responsiveness towards multiple vaccines within the same individuals over the entire adult age-range.

Booster QIV vaccination induced comparable H3N2 hemagglutination inhibition (HI) titers in all age groups, whereas primary PCV13 and mRNA-1273 vaccination induced lower antibody concentrations in older as compared to younger adults. The persistence of humoral responses towards the 6 months timepoint was shorter in older adults for all vaccines. Interestingly, the quantity of vaccine-induced humoral immunity within one individual differed between vaccines. Yet, a small group of mostly older male adults responded low to multiple vaccines.

This study aids the identification of risk groups for low vaccine responsiveness and guides the design of more targeted vaccination strategies for the ageing population.

## Introduction

Vaccination is crucial in our fight against morbidity and mortality caused by infectious diseases. Yet, some individuals, specifically those of older age are not sufficiently protected by today’s vaccination programs, which entails risks for severe disease. These gaps in protection lead to high medical costs and increased societal impact of infectious diseases in the rapidly ageing population^1–3^. Hence, more effective vaccination strategies for older adults are urgently needed.

Currently, annual influenza vaccination as well as vaccination against pneumococci is advised for adults above 65 years of age in most countries. Likewise, older individuals have been a focus group for vaccination against severe acute respiratory syndrome coronavirus 2 (SARS-CoV-2). However, vaccination efficacy as well as humoral and cellular immune responses decline with advancing age^4–9^. This decline in vaccine-induced immunity is a result of a general functional deterioration of the immune system with advancing age, also referred to as immunosenescence^10–13^. Previous research has unraveled phenotypical changes in the ageing immune system, and has identified major influences for non-heritable factors in ageing immunity^14^. Still, this has not yet led to a better understanding of reduced vaccine responsiveness.

Remarkably, vaccine responsiveness is highly variable among individuals, indicating that the pace of immunosenescence varies between individuals^15,16^. Subsequently, future successful vaccination strategies for the ageing population could be directed more towards groups or individuals most at risk for severe infections and low vaccine responsiveness, rather than chronological age only.

The development of new vaccination strategies for these groups is currently hampered by our lack of understanding of the immunological mechanisms underlying inferior responses to vaccination. Moreover, head-to-head comparison of an individuals’ responses towards multiple vaccines is required to determine whether the risk for low vaccine responsiveness depends on vaccine type or is transcending over multiple vaccines. To identify risk groups for low vaccine responsiveness, associations of demographic characteristics and health status with low vaccine responsiveness should be thoroughly investigated. The age-associated decline in health, leading to physical impairment, disease and mortality could be captured in the Frailty Index^17,18^.

To increase our knowledge on vaccine responsiveness with advancing age, we here present the results of an unique vaccination study, in which individuals divided over 3 age groups (young adults 25-49y, middle-aged adults 50-64y, older adults ≥65y) were consecutively vaccinated with 3 different vaccines within a timeframe of two years^19^. Every participant received a seasonal quadrivalent inactivated influenza (QIV) booster vaccination, followed by a primary 13-valent pneumococcal conjugate (PVC13) vaccination up to a year later, and finally a primary vaccination series with the SARS-CoV-2 mRNA-1273 (mRNA-1273) vaccine.

This study primarily aimed to compare the humoral vaccine responses 28 days following primary (PCV13 and mRNA-1273) and booster (QIV) vaccination between the different age groups. Secondary this study aimed to compare the persistence of humoral responses 6 months post-vaccination between the age groups. Finally, this study has the unique opportunity to explore vaccine responsiveness transcending over multiple different vaccines within the same individual and associate responsiveness to health demographics such as the Frailty Index.

Our results indicate comparable induction of humoral immunity following booster QIV vaccination in all age groups, whereas lower humoral responses following primary PCV13 and mRNA-1273 vaccinations were observed in older adults. Secondly, a shorter persistence of humoral responses following all vaccines was observed in the older adults. Importantly, we show that the quantity of vaccine-induced humoral immunity within one individual is vaccine-type specific. However, a small group of majorly older male adults showed low antibody concentrations following multiple vaccines. This finding provides leads to further identify risk groups for low vaccine responsiveness.

## Results

### Study population

A total of 326 participants were included in this study (**Figure 1A**), of whom 315 participants met the per protocol (PP) criteria for at least one vaccine (young adults n = 59, middle-aged adults n = 95, older adults n = 161) (**Figure 1B**). The average age of young, middle-aged and older adults was 36 (range 25-49), 58 (range 50-64), and 76 (range 65-98) years respectively (**Supplementary table 1)**. 34%, 41% and 53% of young, middle-aged and older adults respectively were male. The mean body mass index (BMI) ranged from 24 in the young to 26 in the older adults. Middle-aged and older adults on average received 5 seasonal influenza vaccinations since 2014, whereas this was on average 3 in the young adults. The median frailty index increased from 0.07 (range 0.0 – 0.27) in the young adults to 0.10 (range 0.01 – 0.36) in the middle-aged and 0.18 (range 0.03 – 0.53) in older adults. As expected, the number of medications and incidence of chronic health conditions were higher in the older age groups compared to the young adults.

**Figure 1.**
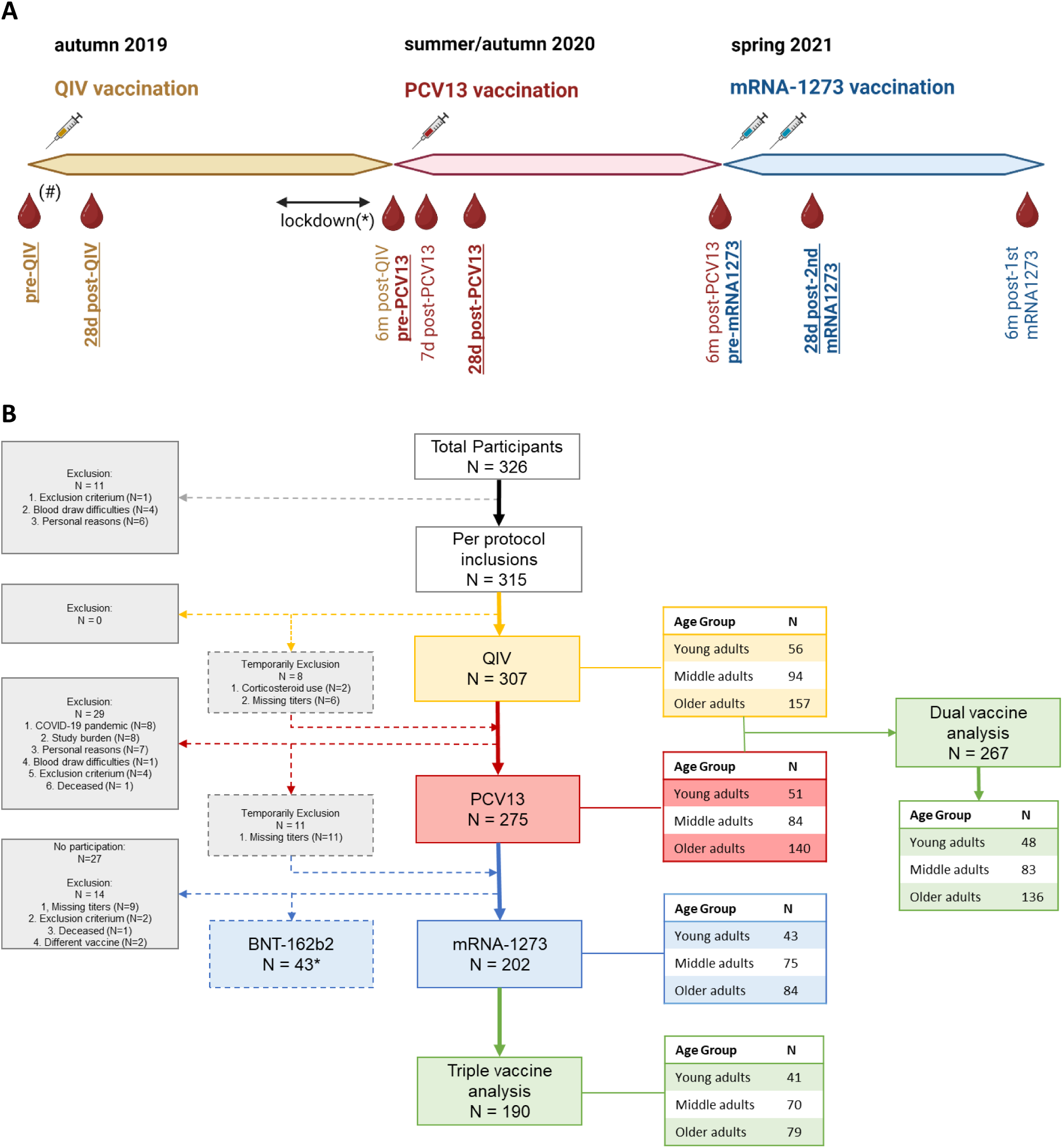
Cohort timeline and participants flow chart. **(A)** Timeline of vaccination cohort with blood drawing timepoints from which samples are used in this reported indicated. The samples used for the primary endpoint analysis (per vaccine) are indicated in bold and underlined. (#) Pre-QIV blood sampling started in summer 2019, while vaccination started in autumn 2019, when the vaccine was available. (*) The SARS-CoV-2 pandemic lockdown caused a temporary interruption of the trial. N=40 participants were sampled before the lockdown. For the other participants, the 6 months post-QIV vaccination time point was extended up to 12 months. Sampling and PCV13 vaccination started again in summer/autumn 2020. **(B)** Participants flow chart indicating the number and age of participants included in the primary endpoint analyses of the different arms of the cohort. Exclusions are divided into definite and temporarily exclusions and reasons for exclusion are given. (*) Large group of older participants were excluded for mRNA-1273 vaccination, due to administration of prior COVID-19 vaccination in the general vaccination program of the Netherlands in winter 2021. This group is analyzed separately as BNT162b2 study group.

The primary endpoint of the study, which is humoral response 28 days post-vaccination, was analyzed in 307 participants (young adults n = 56, middle-aged adults n = 94, older adults n = 157) for the QIV vaccine. In addition, the primary endpoint for PCV13 vaccination was analyzed in 275 participants (young adults n = 51, middle-aged adults n = 84, older adults n = 140) and the primary endpoint for mRNA-1273 in 202 participants (young adults n = 43, middle-aged adults n = 75, older adults n = 84). Importantly, 43 older individuals (mean age = 84y (range 76-98 years)) received BNT126b2 vaccination during the regular COVID-19 vaccination program in the Netherlands and hence were analyzed separately.

Additionally, both QIV and PCV13 vaccine responsive data were available and analyzed in 267 participants (young adults n = 48, middle-aged adults n = 83, older adults n = 136) while 190 participants (young adults n = 41, middle-aged adults n = 70, older adults n = 79) had data available for all three vaccines, including mRNA-1273. The numbers and reasons for exclusions in every step of the clinical trial are depicted on the left side of **Figure 1B**. Of note, some participants were temporally excluded for QIV (n = 8) and PCV13 (n = 11) analysis due to missing titer information or short-term corticosteroid use.

### Booster QIV vaccination induced protective H3N2-specific HI titers in the majority of individuals from all age groups

QIV responsiveness was analyzed by investigating influenza A-specific hemagglutination inhibition (HI). The annual booster QIV vaccination induced a significant (p<0.0001) increase in H3N2 HI titer in all age groups compared to the pre-QIV timepoint (**Figure 2A**). The percentage of H3N2-specific responders, based on an HI titer of ≥40 at 28 days post-QIV vaccination and an increase of >4 as compared to the pre-QIV timepoint, was 61%, 66% and 60% in the young adults, middle-aged adults and older adults respectively. In contrast, an H1N1 HI response was only found in 10% of the participants, based on the same responder criteria (data not shown). Hence, influenza A specific responsiveness following QIV vaccination was further studied using the H3N2 specific HI titers.

**Figure 2.**
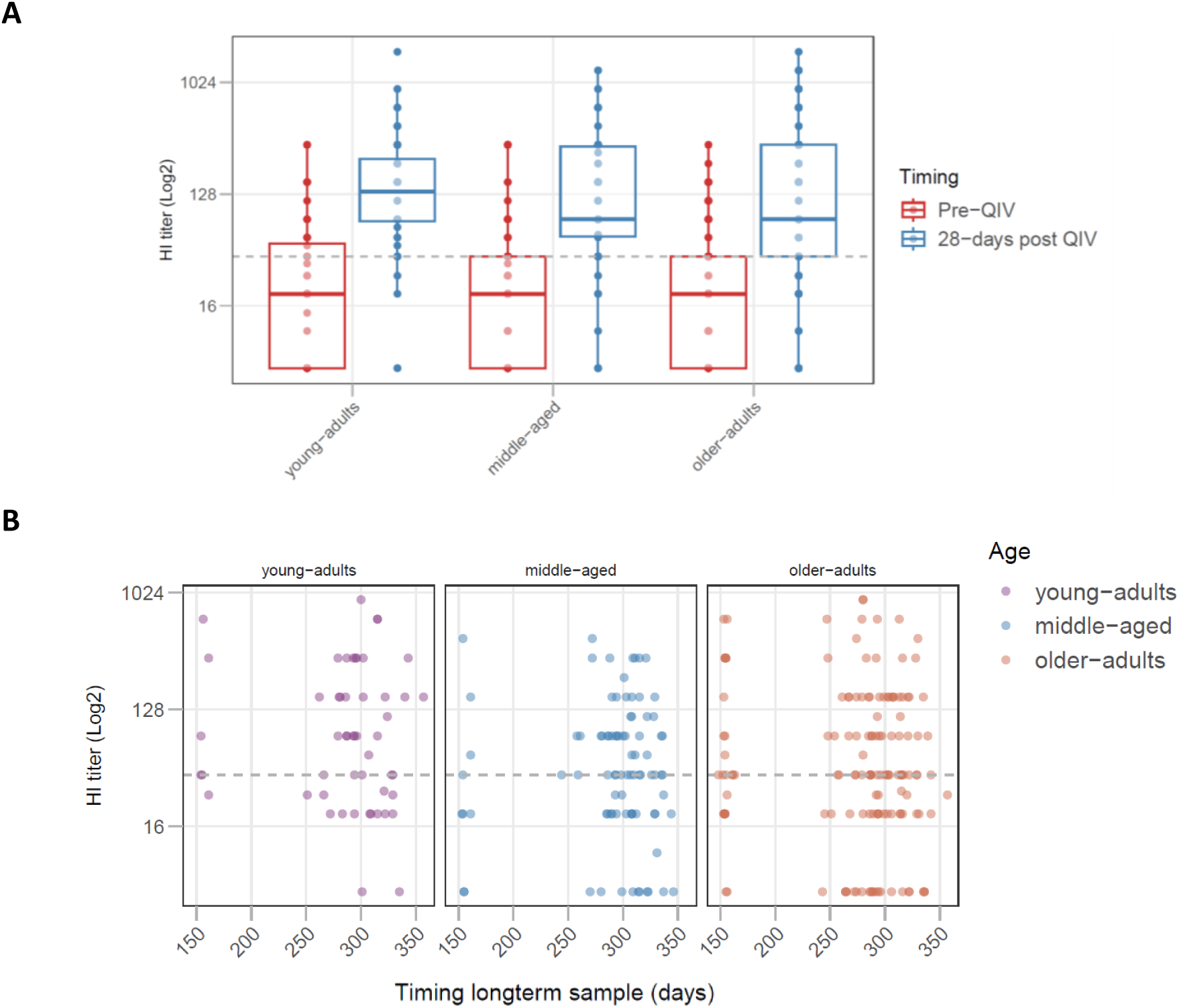
Analysis of QIV vaccination induced H3N2 specific HI titers. **(A)** The pre-QIV and 28-days post QIV vaccination H3N2 specific HI GMT titers split by age group (young adults n = 56, middle-aged adults n = 94, older adults n = 157). The boxplots indicate the median and interquartile range. **(B)** The long-term H3N2 specific HI titers per age groups plotted at the timepoint of sampling (young adults n=52, middle-aged adults n=87, and older adults n=142). In both A and B, the grey dotted line indicates an HI titer of 40, the cut-off for protection.

The 28 days post – QIV booster vaccination H3N2 specific HI titers did not significantly differ between the age groups (median (min – max)): Young adults = 136.5 (5 - 1810), middle-aged adults = 80.0 (5 – 1280), and older adults = 80.0 (5 – 1810)) (**Figure 2A**). Moreover, no correlation between age and H3N2 specific HI titers 28 days post-vaccination was observed (**Supplementary Figure 1A**).

A moderate positive correlation between the pre-vaccination and 28 days post-vaccination H3N2 titers was observed in all age groups (young adults: r= 0.444, middle-aged: r= 0.487, older adults: r= 0.527 (**Supplementary. Figure 1B**)).

Persistence of the humoral response was assessed in 281 participants (52 young adults, 87 middle-aged adults, and 142 older adults). Due to the COVID-19 lockdown, sampling of this timepoint was not uniform between individuals, with the majority of the samples drawn around 300 days post-QIV vaccination in all age groups (**Figure 2B**). Middle-aged and older adults show proportionally more participants with H3N2 HI titers below 40 (young adults = 28.3%, middle-aged adults = 36.8%, and older adults = 36.6%) at this timepoint (**Figure 2B**). More importantly, 3.8%, 13.8%, and 16.2% of the young, middle-aged, and older adults respectively possessed titers under the detection limit of this assay 6 months post-vaccination.

### Primary PCV13 vaccination induced lower IgG responses in older adults

Next, we analyzed the pneumococcal serotype-specific antibodies induced by the primary PCV13 vaccination. A significant increase in IgG concentrations at the 28 days post-vaccination timepoint was observed for all 13 serotypes in all age groups (p<0.0001 for all). Pre-vaccination, older adults had significantly lower IgG concentrations for strain 1, 3 and 6B, whereas middle-aged adults showed significantly higher strain 14-specific IgG as compared to the other age groups (**Figure 3A**).

**Figure 3.**
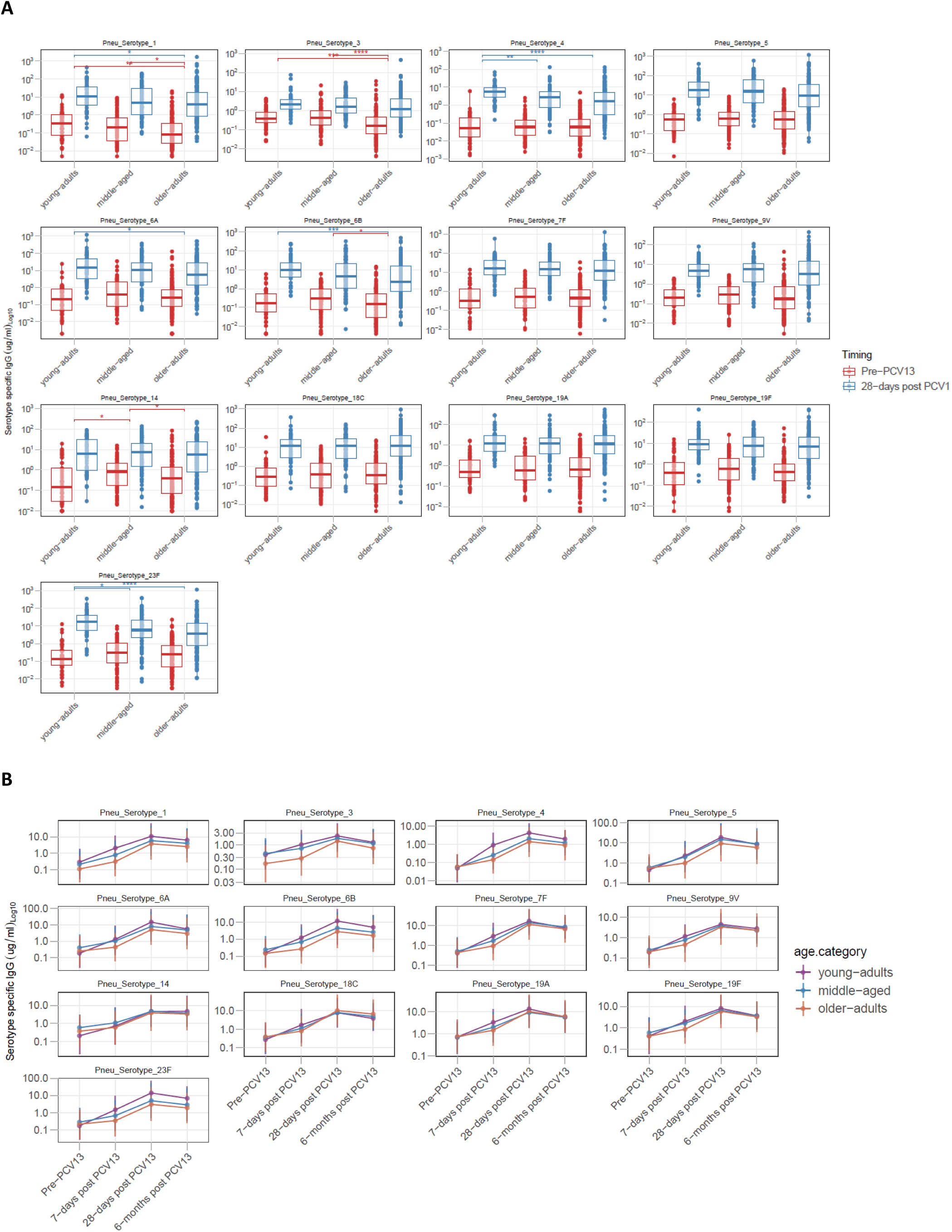
Analysis of PCV13 vaccination induced pneumococcal serotype specific IgG concentrations. **(A)** The pre- and 28 days post-PCV13 vaccination pneumococcal serotype specific IgG concentrations (ug/mL) (presented on a log10 scale) split by age group (young adults n = 51, middle-aged adults n = 84, older adults n = 140). The boxplots indicate the median and interquartile range. *p<0.05, **p<0.01, ***p<0.001, ****p<0.0001. **(B)** The longitudinal (pre-, 7 days, 28 days, and 6 months post PCV12 vaccination) pneumococcal strain specific IgG concentrations split by the different age groups (young adults n= 45, middle-aged adults n= 78, and older adults n= 134). The lines indicate the mean and standard error of the mean.

28 days post-PCV13 vaccination, the IgG concentrations specific for serotypes 1, 4, 6A, 6B, and 23F, were found significantly lower in older adults as compared to the young adult, whereas middle-aged adults also showed a significantly lower serotype 4 and 23F-specific IgG concentration as compared to the young adults (**Figure 3A**). A trend towards a lower IgG concentration at the 28 days post-PCV13 vaccination timepoint was also observed for the remaining serotypes (**Supplementary Table 2**). Weak but significant negative correlations between age and the 28 days post PCV13 IgG concentrations were observed for the pneumococcal serotypes 1,4,5,6A,6B, and 23F (**Supplementary Table 3**).

In addition, positive correlations were observed between the pneumococcal serotype specific pre-vaccination and 28 days post-PCV13 vaccination IgG concentrations for the majority of serotypes. The strength of these correlations was similar in all age groups (**Supplementary Figure 2**).

In all age groups, the IgG concentrations specific for serotypes 5, 6A and 6B strongly correlated amongst each other. Interestingly, the 28 days post-PCV13 vaccination IgG concentrations for the remaining serotypes did not or only weakly correlate within the young adults, while a stronger correlation was observed in the middle-aged and especially older adults (**Supplementary Figure 3**).

PCV13 induced IgG concentrations were longitudinally (pre-PCV13 and 7 days, 28 days, and 6 months post-PCV13) assessed in 45 young adults, 78 middle-aged adults and 134 older adults. 7 days post-PCV13, lower IgG concentrations were observed in older adults for all serotypes, whereas also middle-aged adults had lower IgG concentrations for serotypes 1, 4 and 23F as compared to younger adults (**Figure 3B**). 6 months post-PCV13 vaccination, significantly lower strain 1, 3, 6B and 23F-specific IgG concentrations were observed only in older adults as compared to young adults.

### A primary series of mRNA-1273 vaccination induced lower Spike specific IgG responses in older adults

Next, we analyzed the induction of humoral responses following 2 doses of the mRNA-1273 vaccine. A strong increase in Spike-specific S1 IgG concentration was observed in all age groups (p<0.0001 in all). Pre-vaccination, only 2 young adults, 4 middle-aged and 4 older adults possessed a Spike-specific S1 IgG concentration above the seropositivity level of 10 BAU/mL. 28 days post 2^nd^ mRNA-1273 vaccination, a strong trend to a lower Spike S1-specific IgG concentration was observed in older adults compared to young adults (p = 0.065) (geometric mean IgG concentration (95% CI): young adults = 2587.8 (2203.9 – 3038.5), middle-aged adults = 2217.9 (1874.5 – 2623.3), and older adults = 1832.3 (1513.5 – 2218.3)) (**Figure 4A**). A weak negative correlation between the Spike S1-specific IgG and age was observed (r = -0.165, p-value =0.02)(**Supplementary Figure 5A**).

**Figure 4.**
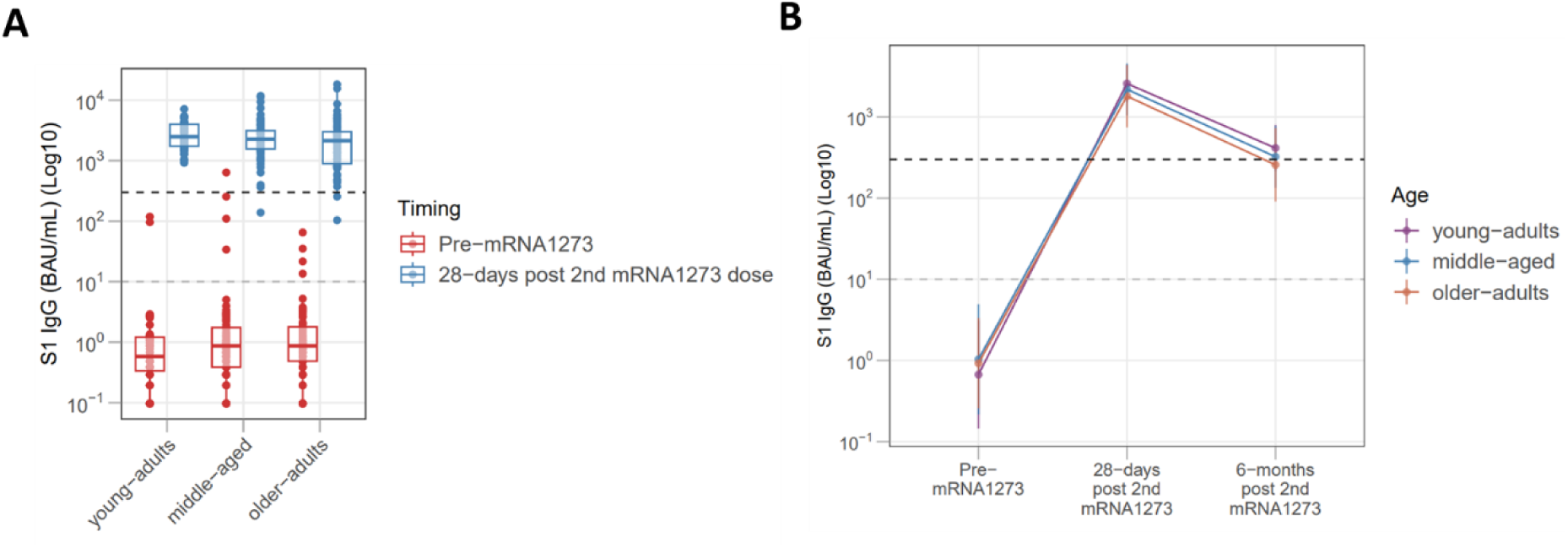
Analysis of mRNA-1273 vaccination induced Spike S1-specific IgG concentrations. **(A)** The pre- and 28-days post 2^nd^ mRNA-1273 vaccination S1 specific IgG concentrations (BAU/mL) (presented on a log10 scale) split by age group (young adults n = 43, middle-aged adults n = 75, older adults n = 84). The box plots indicate the median and interquartile range. **(B)** The longitudinal (pre-, 28 days, and 6 months post 2^nd^ mRNA-1273 vaccination) S1 specific IgG concentrations split by the different age groups (young adults n= 41, middle-aged adults n= 73, and older adults n= 82). The lines indicate the mean and standard error of the mean. No statistics are indicated in this graph. In both A and B, the grey dotted line (S1 specific IgG = 10 BAU/mL) indicates the cut-off for seropositivity. The black dotted line (S1 specific IgG = 300 BAU/mL) indicates the cut-off for a high response.

In addition, 6 months post-2^nd^ mRNA-1273 vaccination, a significantly (p = 0.014) lower S1-specific IgG concentration was observed in older adults as compared to young adults (geometric mean IgG concentration (95% CI): young adults = 414.3 (336.6 – 509.8), middle-aged adults = 323.9 (263.9 – 397.5), and older adults = 257.6 (204.9 – 323.9)) (**Figure 4B**).

Noteworthy, the oldest adults who were vaccinated with BNT162b2 in the general vaccine program (mean age 84 year, range 76-98 years) also showed an adequate (above 300 BAU/mL) geometric mean 629.3 (462.5 – 1036.0) Spike S1-specific IgG concentration 28 days post 2^nd^ BNT162b2 vaccination (**Supplementary Figure 4A**). A trend towards a significant negative association between the Spike S1-specific IgG concentrations and age was also observed in this sub cohort (r = -0.279, p-value = 0.07) (**Supplementary Figure 5B**). In this group, the S1-specific geometric mean IgG concentration at 6 months post 2^nd^ BNT162b2 vaccination was reduced to 88.1 (52.1 – 149.2) (**Supplementary Figure 4B**).

### Low vaccine-induced humoral immunity transcends over multiple vaccines for a small group of mainly older male adults

Finally, an individual’s responsiveness towards multiple vaccines was investigated. Initial analysis indicated an absence of correlation between the 28 days post QIV, PVC13 and mRNA-1273 vaccination antibody titers in all age groups.

Due to the absence of consensus on protective cut-offs and responder profiles for PCV13 vaccination and to allow comparison between the different vaccine-types, a response score was defined. In brief, the 28 days post-QIV and PCV13 antibody titers were divided into quartiles and an individual’s response score was determined for both vaccines separately (1 = low, 2-3 nominal, and 4 = high) (for more details see methods section). Subsequently response scores for the 2 vaccines were compared per individual and visualized in **Figure 5A**. This analysis reveals large differences between QIV and PCV13 induced humoral immunity within one individual. The percentage of individuals in the lowest category (1) was lowest in the young adults (QIV 14.6% and PCV13 8.3%) and largest in the older adults (QIV 30.1% and PCV13 30.9%) (**Supplementary Table 4**). Interestingly, the proportions of individuals in the highest response category (4) was similar between the young (QIV 27.1% and PCV13 25%) and older adults (QIV 25.7% and PCV13 29.4%). The dual vaccine response score per individual was defined as the average score between QIV and PCV13. 0% of young adults, 6% middle-aged adults and 11% older adults were found in the lowest dual response category (1), whereas this division is 29.2%, 18.1% and 17.6% for the highest dual response category (4) respectively.

**Figure 5.**
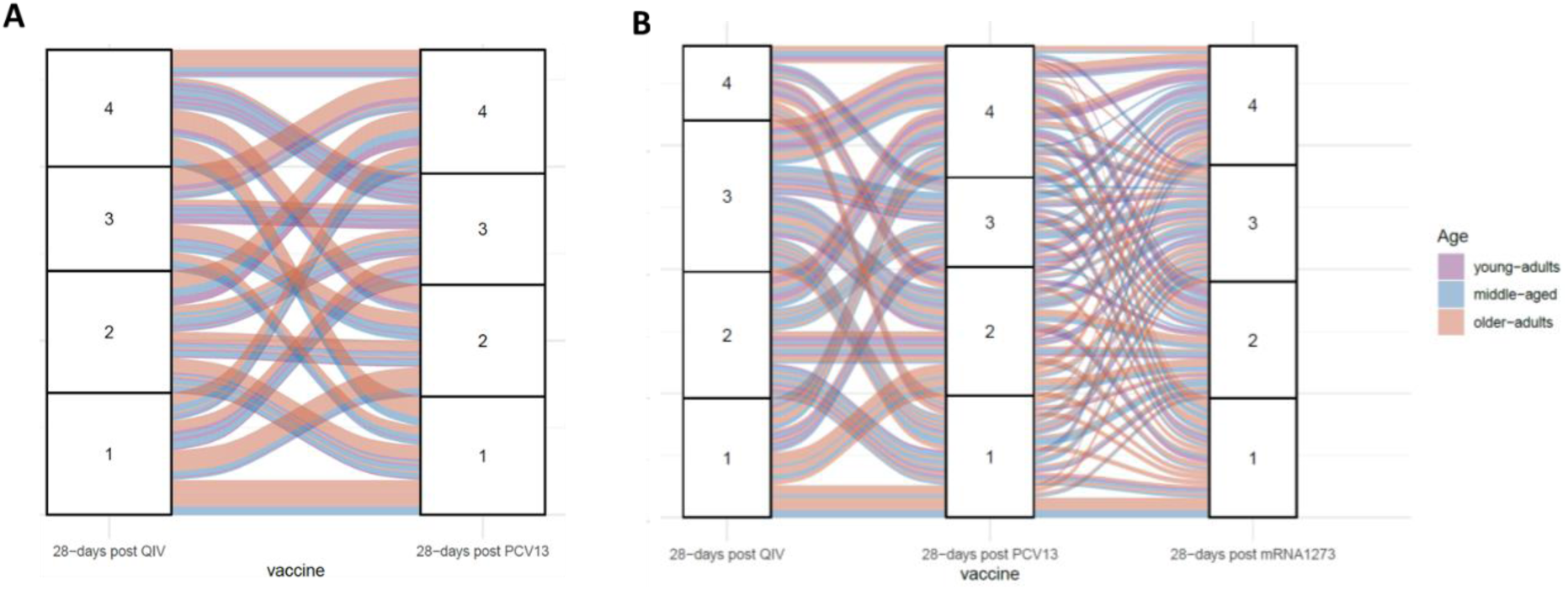
Multiple vaccine responsiveness. **(A)** Dual (QIV and PCV13) and **(B)** Triple (QIV, PCV13, and mRNA-1273) vaccine responsiveness. The 28 days post-vaccination antibody titers are divided into quartiles, where number 1 indicated the lowest and number 4 the highest quartile of responders. Per vaccine, each individual is assigned to a quartile based on the 28 days post-vaccine antibody concentration. Every lined indicate the trajectory of an individual between the different vaccines indicated. The line color indicated the age group of an individual. In **(A)**: young adults n= 48, middle-aged adults n= 83, and older adults n= 136 and in **(B)** young adults n= 41, middle-aged adults n= 70, and older adults n= 79. In B a large group of older adults is excluded based on the administration of the BNT162b2 vaccine in the general COVID-19 Dutch vaccination program, which changes the quartile division.

Strikingly, 17 participants (5 middle-aged adults and 12 older adults) were assigned the highest score for QIV but the lowest for PCV13. Likewise, 16 participants (1 young adult, 7 middle-aged adults and 8 older adults) scored lowest for QIV and highest for PCV13. In total, 20 (7.5%) participants scored low and 53 (19.9%) participants high for both vaccines. In addition, 60 (22.5%) participants responded nominal (responds score 2 and 3) for both vaccines.

Next, we compared the demographic characteristics of individuals in the 4 different dual response groups (**Table 1**). Individuals in the lowest dual vaccine response group were older as compared to the other response groups and more often of the male sex. In addition, a lower pre-vaccination H3N2 vaccination titer was observed in the lowest dual vaccine response group, whereas BMI, the Frailty Index, EQ-5D-3L score, and the number of prescription medications used was equal between the groups. Even though highly spread, the highest mean pre-vaccination H3N2-specific HI titers were found in the highest dual vaccine response group.

**Table 1.** Demographic characteristics of dual and triple vaccine response groups.

| Dual vaccine response group |  |  |  |  |
| --- | --- | --- | --- | --- |
|  | 1<br>n = 20 | 2<br>n = 80 | 3<br>n = 114 | 4<br>n = 53 |
| **Age**<br>mean (min-max) | 71.9 (58-89)<sup>*c</sup> | 63.5 (25 – 92) | 62.2 (25 – 90) | 60.8 (25 – 89) |
| **% Males** | 70<sup>*a, *b</sup> | 40 | 46 | 55 |
| **BMI**<br>mean (min-max) | 26.6 (20.1 – 40.6) | 25.2 (18.7 – 37.9) | 25.3 (17.4 – 37) | 25.1 (19.6 – 33.0) |
| **H3N2 pre-**<br>**vaccination titer**<br>median (min-max) | 5 (5 – 40)<sup>**b, ****c</sup> | 7.5 (5 – 80)<sup>****c</sup> | 20 (5 – 320)<sup>**c</sup> | 40 (5 – 320) |
| **Frailty index**<br>median (min-max) | 0.15 (0.01 – 0.53) | 0.12 (0.0 – 0.45) | 0.14 (0.0 – 0.43) | 0.12 (0.0 – 0.40) |
| **EQ-5D-3L**<br>median (min-max) | 0.87 (0.40 – 1) | 1 (0 – 1) | 1 (0.17 – 1) | 0.84 (0.33 – 1) |
| **Number of**<br>**medications**<br>median (min-max) | 3 (0 -11) | 1 (0 – 12) | 2 (0 – 9) | 2 (0 – 11) |
| Triple vaccine response group |  |  |  |  |
|  | 1<br>n = 17 | 2<br>n = 82 | 3<br>n = 77 | 4<br>n = 14 |
| **Age**<br>mean (min-max) | 65.9 (41 – 78) | 60.1 (25 – 78) | 56.7 (26 – 78) | 54.6 (30 – 73) |
| **% Males (Range)** | 82<sup>**a, **b,</sup> | 44 | 43 | 64 |
| **BMI**<br>mean (min-max) | 26.2 (20.1 – 37) | 25.1 (17.4 – 37.9) | 25.2 (19.6 – 38.8) | 25.6 (21.3 – 33.0) |
| **H3N2 pre-**<br>**vaccination titer**<br>median (min-max) | 5 (5 – 80)<sup>*c</sup> | 20 (5 – 226) | 20 (5 – 320) | 40 (5 – 320) |
| **Frailty index**<br>median (min-max) | 0.15 (0.03 – 0.31) | 0.11 (0.0 – 0.39) | 0.11 (0.0 – 0.31) | 0.15 (0.0 – 0.36) |
| **EQ-5D-3L**<br>median (min-max) | 1 (0.78 – 1) | 1 (0.30 – 1) | 1 (0.30 – 1) | 0.84 (0.33 – 1) |
| **Number of**<br>**medications**<br>median (min-max) | 3 (1 - 7) | 1 (0 – 10) | 1 (0 – 9) | 2.5 (0 – 1) |

<sup>a</sup>: compared to group 2. <sup>b</sup>: compared to group 3. <sup>c</sup>: compared to group 4. \*p<0.05, \*\*p<0.01, \*\*\*p<0.001, \*\*\*\*p<0.0001. Group 1 represents individuals with an overall vaccine response score of 1; low responders. Groups 2 and 3 consist of individuals with a nominal responds, and group 4 represents the group of individuals with a high overall vaccine response score.

Furthermore, analysis of triple vaccine responsiveness (**Figure 5B**) in a sub cohort of 190 individuals (see **Figure 1B**) indicated a large mixture in humoral immunity induced by multiple vaccines, both within and between individuals. Due to the different group size in this analysis, quartiles in antibody concentrations slightly changed. Yet again, the percentage of individuals in the lowest category was smallest in the young adults (1) (QIV 12.2%, PCV13 12.2%, mRNA-1273 12.2%) and largest in the older adults (QIV 34.2%, PCV13 31.6%, mRNA-1273 35.4%). The proportions of individuals in the highest response category (4) was slightly lower in the older adults (QIV 17.7%, PCV13 27.8%, mRNA-1273 22.8%) as compared to the young (QIV 22.0%, PCV13 34.1%, mRNA-1273 34.1%).

There are several extremes in this triple vaccine analysis: 2 older adults scored high for QIV vaccination, while low for both PCV13 and mRNA1273. In addition, 5 participants (2 middle-aged adults, and 3 older adults) scored high for PCV13 while low for QIV and mRNA1273. Only 1 older adult scored high for mRNA-1273, while low for QIV and PCV13.

In this analysis, a group of 17 (8.9%) and 14 (7.4%) individuals was designated as overall low and high responders respectively, while 26 (13.7%) participants responded nominal for all three vaccines. Yet again, the lowest triple vaccine responders were of higher age and were mainly of the male sex, whereas frailty status and BMI did not differ (**Table 1**). Compared to the highest response group, the lowest responders had significantly lower pre-vaccination H3N2 titers.

Combined these results indicate that the levels of vaccine-induced humoral immunity is highly vaccine specific, though low responsiveness is transcending over multiple vaccines for a small group of mainly older male adults.

## Discussion

The primary outcomes of this unique clinical trial indicate equal peak antibody responses following an annual booster QIV (season 2019-2020) vaccination in old and young individuals, while lower humoral responses are observed following both primary PCV13 and mRNA-1273 vaccinations in older adults compared to middle-aged and young adults. Additionally, our results suggest a shorter persistence of this humoral immunity in older adults for all vaccines. Importantly, the analysis demonstrates vaccine-specific humoral responsiveness, although a small group of mainly older male adults is classified as low responders to multiple vaccines.

Contrary to previous indications of reduced responsiveness to annual influenza vaccination in older adults ^9,20–23^, the 2019-2020 booster QIV vaccine administered in this cohort induced statistically equal H3N2-specific HI responses 28 days post vaccination in young, middle-aged and older adults, including an equal proportion of responders. One explanation for this could be potential differences in immunogenicity between seasonal influenza vaccinations and strains. Moreover, most existing studies on ageing and influenza vaccine responsiveness derive from cohorts that exhibit substantial differences in pre-vaccination immunity between younger and older individuals. By ensuring the administration of a booster vaccination in all age groups, based on the inclusion of previously influenza vaccinated individuals in 2018-2019, the current study partly mitigates the impact of pre-vaccination differences in the analysis of QIV responsiveness. While our analyses indeed indicate a substantial association between the pre-vaccination and 28 days post-vaccination H3N2-specific HI titers, no difference in pre-vaccination H3N2-specific HI titers was observed between the age groups.

It is however of interest to investigate whether the kinetics of the recall response is delayed in older adults.

Pre-existing immunity is not only reflected in the presence of pre-vaccination HI titers, but also influenza-specific cross-reactive T-cells are of importance^24^. To fully understand the effect of pre-existing immunity on subsequent QIV responsiveness, future research should additionally investigate QIV-specific T-cell immunity. This investigation is of special relevance due to the currently acknowledged large impact of ageing on the T-cell compartment^25^.

Remarkably, low H1N1-specific HI responses were observed after the QIV vaccination, as also previously noted by others^26^, which limited our abilities to investigate differences in humoral H1N1-specific vaccine responses between age groups. We suggest that seasonal strain-specific effects and limited antigenic drift between vaccine strains in previous years, resulted in high pre-vaccination H1N1-specific immunity and restricted the subsequent H1N1-specific immune response to the vaccination.

The lower humoral responsiveness following the primary PCV13 and mRNA-1273 vaccinations in older adults fits with the general concept of reduced potential to respond to *de novo* antigens at older age^27^ and potentially relates to a general decline of naïve immune cells with advancing age^28–30^. In line with this, lower responses following primary vaccinations, such as SARS-CoV-2^5,31^, pneumococci^32,33^, yellow fever^34^, and Japanese encephalitis^35^ vaccinations in older adults were previously observed by others. Nevertheless, comparison of primary vaccine responses between younger and older adults is often complicated by a difference in pre-vaccination immunity induced by historical natural exposures. Although PCV13 vaccination was a primary challenge in this cohort, previous natural exposure is evidenced by a large variation in serotype-specific pre-vaccination IgG concentrations between individuals and expected due to high natural circulation of pneumococci^36^. These previous exposures likely influenced pre-vaccination humoral and cellular immunity.

The observation of a slower induction of PCV13-specific antibodies in older adults either indicates a delay in immune responses at older age, as previous also observed following yellow fever vaccination^34^, or indicates lower pre-vaccination immunity and hence an immune response more closely resembling a primary response in older adults. The latter is however contraindicated by the presence of a moderate correlation between pre- and post-PCV13 vaccination serotype specific IgG responses in all age groups. Despite this finding, the stronger correlation between serotype-specific IgG responses in older adults as compared to younger age groups warrants further investigation into difference in cellular pre-vaccination immunity between the age groups. Specifically, in-depth investigation of pneumococcal-specific cellular immunity, but also immunity against the conjugate of the vaccine, CRM197, in the different age groups potentially helps to unravel the mechanisms underlying the deviating immune responses observed to PCV13 vaccination in older adults.

The SARS-CoV-2 vaccination campaign provided a unique opportunity to investigate an additional primary vaccine response in relation to age. Despite adequate vaccine responses in all age groups, a strong trend towards a lower peak antibody response was observed in the older adults, which is in agreement with previous findings^5^. Noteworthy, exclusion of the oldest individuals, due to the administration of a primary series of BNT162b2 vaccination to these individuals in the general vaccination campaign of the Netherlands preceding our vaccination initiatives, likely impacted the statistical power of this comparison.

Besides lower peak antibody responses to PCV13 and mRNA-1273 vaccination, our results might also indicate a lower persistence of humoral responses in older adults following all three vaccinations. This may be due to a decline in the survival of long-lived plasma cells with advancing age and is in accordance with the general understanding of decreased survival niches for long-lived plasma cells in the ageing bone marrow^37^. Secondly, this observation is indicative of a diminished memory B cells response, potentially due to the involvement of Age Associated B (AAB) cells^38^, following vaccination in older age groups, as was also previously noted ^39,40^. Therefore, booster vaccinations might be needed at older age in order to maintain long-term protection.

The large variation in antibody titers induced by the different vaccinations in the older age groups underlines the currently acknowledged deviating pace of immunosenescence between ageing adults^15^, and hence supports the use of risk profiles, instead of chronological age, in the design of future vaccination strategies for the ageing population. Early signs of ageing immunity are already visible in the middle-aged adults group, as evidenced by a large variation in vaccine responsiveness and a significantly lower PCV13 response for two pneumococcal serotypes as compared to the young adults. In line, lower vaccine responses towards a primary meningococcal vaccine were previously found in middle-aged adults as compared to adolescence ^41^. Nevertheless, only a few middle-aged adults were found amongst the lowest dual and triple vaccine responders, indicating induction of effective responses by all vaccine types in this age group. Hence middle-aged adults are an interesting target for future vaccine strategies, in order to strengthen memory immunity in the general population before reaching older age ^42,43^.

Importantly, the unique setup of the presented clinical trial allowed investigation of vaccine responsiveness, including risk group identification for low responsiveness, transcending over multiple vaccines. This novel head-to-head comparison of humoral immunity to multiple vaccines within the same individual, reveals that the level of vaccine-induced humoral immunity is highly vaccine specific. The contradictory classification of some individuals as high responders following QIV vaccination and at the same time low responders following PCV13 and mRNA-1273 vaccination, indicates that this vaccine-specific responsiveness is partly explained by the nature of the induced immune response; either a booster or a primary response. Combined with the observed higher pre-vaccination H3N2 HI titers in the highest response groups, this suggests an important influence of pre-vaccination immunity in the vaccine response at older age. Moreover, the effectiveness of the different vaccine platforms used might differ between individuals, where mRNA vaccines suggest to induce stronger humoral responses in older individuals^44^ and conjugate vaccine were found more effective as compared to plain polysaccharide vaccines^45^.

Yet, a small group of predominantly older male individuals responded low to all vaccines. Previously, sex has been described as an important parameter in vaccine responsiveness^46^ as well as immune ageing^47^. Moreover, a faster pace of immunological ageing has been suggested in older males^48,49^.

Contrary to previous research that observed a correlation between the Frailty Index and humoral responses to SARS-COV2 vaccination in older adults^50^, no difference in frailty status was observed between the response groups in this cohort. The size of our cohort and the relatively healthy status of the participants, in which nursing home residents were excluded, are factors likely affecting our analysis. Yet, it is of interest to investigate whether parameters more closely resembling the immune status have a higher predictive value for vaccine responsiveness.

Besides its unique set-up, this study has several limitations. Firstly, the absence of validated correlates of protection (CoPs) for pneumococci in older adults complicated the analysis of multiple vaccine responsiveness. Subsequently the scoring methods used did not allow identification of non-responders, but instead identifies individuals with low vaccine-induced humoral immunity. Secondly, as a result of the COVID-19 lockdowns, blood sampling timepoints have been postponed, limiting the ability to analyze the long-term response following QIV vaccination. Moreover, the young and middle-aged adults included in this cohort are mostly health care workers, and might not fully represent the general population. Finally, the current analysis is performed on antibody concentrations. Despite previously observed correlations between antibody concentrations and vaccine efficacy^51,52^, it is of interest to also investigate antibody functionality in relation to ageing.

Therefore, future research should investigate whether ageing potentially affects antibody quality to a higher degree as compared to quantity.

Taken together, the outcomes indicate a potential large impact of pre-vaccination immunity on vaccine responsiveness at older age. Moreover, the presented study accommodates the identification of risk groups for low vaccine responsiveness and provides leads for further research to untangle mechanisms underlying these low responses. Hence, this study supports development of more targeted vaccination strategies for the rapidly ageing population.

## Methods

### Study design and participants

We here report on the longitudinal intervention studies VITAL and VITAL-corona^53^. Samples of these primary endpoints were collected between 2019 and 2021. Within this cohort, participants divided over 3 age groups were recruited: young adults (25-49y), middle-aged adults (50-64y) and older adults (≥65y). The young and middle-aged adults were recruited among workers of public healthy institutions of the University Science Park and University medical center Utrecht, The Netherlands. Older adults were recruited from a previous cohort^54,55^. All participants needed to be capacitated and vaccinated with the seasonal influenza vaccination in season 2018-2019 to be considered eligible for participation. At the start of the intervention cohort (autumn 2019), potential participants were excluded based on the following criteria: received a previous pneumococcal vaccination, known or suspected allergy to any of the vaccine components, received a high systemic (>20 mg) daily dose of corticosteroids within 2 weeks before inclusion, use of high (>30 mg) dose of corticosteroids in the recent past, recipient of an organ or bone marrow transplant, have a (functional) asplenia, received chemotherapy in the past 3 years, received blood products or immunoglobulins within 3 months before inclusion, known or suspected coagulation disorder that would contraindicate against intramuscular injection and frequent blood sampling, known or suspected immunodeficiency or use of immunosuppressive therapy, known anemia, or known infection with immunodeficiency virus (HIV) and/or hepatitis B and/or C virus. Participants were additionally excluded for the mRNA-1273 vaccination when having received treatment with COVID-19 monoclonal antibodies within 3 months before vaccination. Moreover, participants were temporarily excluded from the study when they: received any vaccine within 1 month of a vaccination visit or within 2 weeks of blood collection. Study visits were postponed when participants were experiencing an elevated body temperature >38 °C within 72 hours before a vaccination visit or 48 hours before a blood collection visit as well as in case of a positive COVID-19 test (visit postponed for at least 4 weeks). Finally, participants were withdrawn from the study when: receiving a systemic high (>20 mg) dose of corticosteroids, starting chemotherapy treatment, receiving blood products or immunoglobulins, being pregnant at the moment of pneumococcal or mRNA-1273 vaccination, or perceiving sudden anemia.

Ethical approval was obtained through the Medical Research Ethics Committee Utrecht (NL69701.041.19, EudraCT: 2019-000836-24). All participants provided written informed consent and all procedures were performed with Good Clinical Practice and in accordance with the principles of the Declaration of Helsinki.

### Vaccinations and blood sampling

A schematic outline of the study design is depicted in **Figure 1**. At the start of the study, a blood sample was collected from all participants (pre-QIV; start summer 2019). Subsequently, all participants were vaccinated with Influvac Tetra (2019-2020) (autumn 2019); the seasonal quadrivalent inactivated subunit influenza vaccine (QIV)(2019-2020), containing neuraminidase and hemagglutinin from the following viral strains: A/Brisbane/02/2018, IVR-190(H1N1); A/Kansas/14/2017, NYMC X-327 (H3N2); B/Maryland/15/2016, NYMC BX-69A (B/Victoria/2/87 lineage); and B/Phuket/3073, wildtype (B/Yamagata/16/88 lineage) (Abbott Biologicals B.V. The Netherlands).

Secondly, during the summer/autumn of 2020, all participants were vaccinated with Prevenar 13, the 13 valent pneumococcal polysaccharide conjugate vaccine containing polysaccharides from the pneumococcal serotypes -1, -3, -4, -5, -6A, -6B, -7F, -9V, -14, -18C, -19A, -19F and -23F conjugated to the CRM197 carrier protein (Pfizer Europe, Belgium).

Finally, during spring 2021, participants received a primary vaccination series with one-month interval with Spikevax, the SARS-COV2 mRNA-1273 vaccine (Moderna Biotech, Spain), unless they already had been vaccinated through the national vaccination program with a BNT162b2 vaccine.

Following each vaccination blood samples were collected 28 days (+/- 3 days) and 6 months (range 5-8 months) post-vaccination. Here the 6 months timepoints also serve as pre-vaccination sample before either PCV13 or mRNA-1273 vaccination. In addition, a blood sample was collected 7 days (+/- 1 day) following PCV13 vaccination and at the moment of second mRNA-1273 vaccination. Of note, in spring 2020, the SARS-COV2 pandemic hit, resulting in a temporary shutdown of the cohort and restart 4 months later. Therefore, the 6 months (window 5-8 months) post-QIV vaccination sampling was extended to a window of 12 months.

Blood samples were collected by venipuncture using blood collection tubes containing clot activator and gel separator (Greiner Bio-one, Austria). Serum was collected and aliquoted within 8 hours after sampling and stored at -80 °C until further use.

### Serological analysis

The humoral response towards the influenza A strains A/Brisbane/02/2018, IVR-190(H1N1) and A/Kansas/14/2017, NYMC X-327 (H3N2) strain at the pre- and post-vaccination timepoints were used to evaluate the response towards the QIV vaccination. The H1N1 and H3N2-specific antibody responses were respectively measured at Vismederi (Siena, Italy) and Viroclinics (Rotterdam, the Netherlands) using the Hemagglutination Inhibition (HI) assay, the most commonly used assays for measuring influenza specific antibody titers, according to the standard methods of the World Health Organization (WHO) as explained in^56–58^. In brief, a dilution series of serum samples was incubated with Hemagglutinin Units (HAU) influenza virus and thereafter incubated with turkey erythrocytes.

Subsequently, agglutination of red blood cells was scored and the antibody titer preventing agglutination calculated. An HI titer >40 was considered protective. A response to the QIV vaccine was define as an HI titer >40 at 28 days post-vaccination and a fold-increase of >4 compared to the pre-QIV timepoint.

The pneumococcal serotype-specific IgG concentrations for the 13 serotypes present in the PCV13 vaccine were measured using the fluorescent-bead-based-multiplex immunoassay (MIA) as previously described in ^59,60^ with minor modifications of using a protein-free buffer (Surmodics) with 10% FCS in the assay. The WHO international standard 007sp was used as a standard. For each sample, median fluorescent intensity was converted to IgG concentration (μg/ml) by interpolation from a 5-parameter logistic standard curve. Results were obtained using a Bio-plex 200 system with Bio-plex software (version 6.2, Bio-Rad, UK).

The Spike S1-specific IgG concentrations induced by the mRNA-1273 vaccine were measured using bead-based assay as previously described^61^. Here the S1-specific concentrations were calibrated against the SARS-CoV2 specific international standard (20/136 NIBSC standard) and expressed as binding antibody units/mL (BAU/mL) and a concentration of 10.1 BAU/mL was used a cut-off for seropositivity.

### Calculation vaccine response profile

In order to define an individual’s vaccine responsiveness towards multiple vaccines, an individual vaccine response score was defined per vaccine.

In order to do so, the 28 days post-vaccination antibody titers per vaccine antigen (QIV: the H3N2 titer, PCV13: the IgG concentrations against the 13 pneumococcal serotypes, and mRNA-1273: the IgG concentrations against the Spike S1 protein) were divided into quartiles. Subsequently, a score of 1 was given to an individual with an antibody titer in the lowest quartile, whereas a score of 4 was given to an individual with an antibody titer in the highest quartile (and 2 and 3 to the middle quartiles).

Since for the PCV13 vaccine, antibody responses were measured for 13 different strains separately, we first defined the response score per serotype. Thereafter we used a majority votes approach (most frequent score among the 13 serotypes), to define the most frequent response score among the 13 serotypes and used this most frequent score as the overall vaccine response score. To break ties for a few cases, where individuals had similar frequency of scores, we randomized the score selection using the mclust package majority Vote function in R.

An individual’s dual or triple vaccine response score was defined as the average between the scores to the QIV and PCV13 (dual) or QIV, PCV13 and mRNA1273 (triple), respectively. Of note, since the triple vaccine response score was investigated in a smaller group of individuals, the quartile division slightly differed between the dual and triple vaccine analyses.

### Frailty status determinants

The Frailty Index, EQ-5D-3L questionnaire score and number of prescription medications were used to assess the frailty status of the participants and were compared between the dual and triple vaccine response groups. The Frailty Index and EQ-5D-3L scores in the VITAL cohort have been described previously^62^. In brief, the Frailty Index is based on 31 deficits and the scores ranged from 0 (least frail) to 0.53 (most frail) in this cohort. The EQ-5D-3L index and number of prescription medications are based on self-reported questionnaires. Answers to the EQ-5D-3L questionnaire were converted using the Dutch population norms, resulting in scores ranging from 1 (least frail) to -.03 (most frail) in this cohort. The number of medications included all medication prescribed by a physician.

### Statistics

The distribution of data was tested before downstream analysis. Age groups were compared at the pre- and 28 days post-vaccination timepoints using the Kruskall-Wallis test and corrected for multiple comparisons with Bonferroni correction.

When comparing paired samples from individuals at two different timepoints, we used the Wilcoxon signed rank test.

The mean (QIV) and geometric mean (PCV13 and mRNA-1273) titers and 95% Confidence intervals (CI) per age group were calculated using the DescTools package, using 999 bootstrap replicates.

Correlations between the 28 days post-vaccination antibody titers were calculated using spearman correlation. Comparison of age, BMI and H3N2 pre-titers between the dual and triple vaccine response groups were performed using the Kruskall-Wallis test and corrected for multiple comparisons with Bonferroni correction. Sex was compared between the vaccine response groups with the Chi-Squared test. In all analyses, a p-values < 0.05 was considered significant. Data handling and visualization was done with tidyverse R tools.

## Data Availability

The datasets generated during the current studies are not made available in a public database since the study is ongoing. We will share pseudonymized data upon reasonable request as long as data transfer is in agreement with the clinical protocol and EU legislation on the General Data Protection Regulation, which should be regulated in a data sharing agreement.

## Acknowledgement

We would like to thank all collaborators; Inge Pronk, Linde Woudstra, Jacqueline Zonneveld, Sandra Hoogkamer, Marjolein Izeboud, Olga de Bruin, Helma Lith, Joyce Geeber, Ilse Akkerman, Megan Barnes, Lysanne Bakker, Shirley Man, Silvia Cohen, Ruben Wiegmans, Nazela Mir Leibady, Ronith van de Wiele, Rydianne Carvalho De Moura, Emma van Wijlen, Petra Molenaar, Madelène Paets, Martien Poelen, Martijn Vos, Jeroen Hoeboer, Marion Hendriks, Noortje Smits, Jolanda Kool, Maarten Emmelot, Ronald Jacobi, Stefanie Lenz, Floor Peters, Gaby Smits and Marjan Kuijer.

## Author contributions

LB, MvH, NR, WB, JvB and DvB designed the study, wrote the medical ethical application and performed the clinical trial. LB, IT, RvB, TO, AB and JvB were involved in data acquisition. EB was responsible for the data management. MvdH, SS, EB, AC, YvS and JvB analyzed the data. MvdH, JvB and DvB wrote the manuscript. All authors critically revised the manuscript before publication.

