## Supplementary material for "Multiple vaccine comparison in the same adults from the VITAL study reveals vaccine-specific and age-related humoral response patterns"

**Supplementary Table 1. Demographics characteristics of study population.**

| | **Young adults** | **Middle-aged adults** | **Older adults** |
|---|---|---|---|
| **Number** | 59 | 95 | 161 |
| **Age -** mean [min-max] | 36 [25-49] | 58 [50-64] | 76 [65-98] |
| **% male** | 34% | 41% | 53% |
| **BMI -** mean [min-max] | 24 [18.7-35.5] | 24.6 [18.7-37.0] | 26.0 [17.4-40.6] |
| **Number of seasonal influenza vaccinations since 2014 -** median [min-max] | 3 [1-5] | 5 [1-5] | 5 [1-5] |
| **Frailty index –** median [min – max] | 0.07 [0.0 – 0.27] | 0.10 [0.01 – 0.36] | 0.18 [0.03 – 0.53] |
| **EQ-5D-3L –** median [min – max] | 1 [0.18 – 1] | 1 [-0.03 – 1] | 1 [0.30 – 1] |
| **Number of medications –** median [min – max] | 0 [0 – 7] | 1 [0 – 7] | 4 [0 – 12] |
| **Cardiovascular disease -** n(%) | 0 (0%) | 4 (4.2%) | 42 (26.1%) |
| **High blood pressure -** n(%) | 4 (6.8%) | 28 (29.5%) | 82 (50.9%) |
| **Cancer -** n(%) | 0 (0%) | 9 (9.5%) | 41 (25.5%) |
| **Diabetes -** n(%) | 0 (0%) | 6 (6.3%) | 20 (12.4%) |
| **Lung diseases -** n(%) | 1 (1.7%) | 7 (7.4%) | 6 (3.7%) |
| **Joint inflammation -** n(%) | 0 (0%) | 3 (3.2%) | 11 (6.8%) |
| **Disease of Nervous system -** n(%) | 0 (0%) | 0 (0%) | 5 (3.1%) |

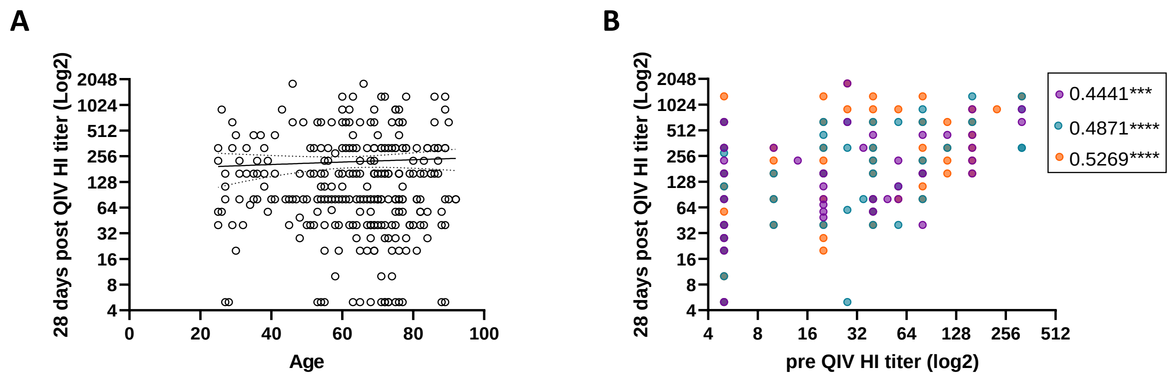


**Supplementary Figure 1. Additional analysis on H3N2 HI titers 28 days post QIV vaccination.**

**(A)** Correlation between the 28 days post-QIV H3N2 HI titers and age**. (B)** The correlation between the pre-vaccination and 28 days post QIV vaccination HI titers per age group (young adults: purple, middle-aged adults: blue, and older adults: orange). The Spearman rho is provided. *** p<0.001, **** p<0.0001.

**Supplementary Table 2. 28 days post PCV13 vaccination pneumococcal serotype specific geometric mean IgG concentrations.**

| *Pneumococcal serotype* | *Young adults* | *Middle-aged adults* | *Older adults* |
|---|---|---|---|
| *1* | 9.80 [5.78-16.61] | 5.56 [3.60-8.59] | 4.11 [2.84-5.93]*[a] |
| *3* | 2.25 [1.64-3.08] | 1.82 [1.39-2.38] | 1.48 [1.14-1.93] |
| *4* | 4.64 [3.32-6.48] | 2.07 [1.46-2.95]**[a] | 1.37 [1.00-1.88]****[a] |
| *5* | 17.67 [11.22-27.81] | 14.53 [9.94-21.24] | 9.18 [6.56-12.83] |
| *6A* | 13.35 [8.03-22.20] | 7.49 [4.79-11.71] | 5.19 [3.66-7.36]*[a] |
| *6B* | 9.99 [6.30-15.83] | 4.46 [2.80-7.12] | 2.77 [1.93-4.00]***[a] |
| *7F* | 16.16 [11.10-23.54] | 13.59 [10.18-18.14] | 11.62 [8.66-15.59] |
| *9V* | 4.75 [3.41-6.62] | 3.92 [2.81-5.48] | 3.30 [2.36-4.62] |
| *14* | 4.84 [2.79-8.40] | 4.73 [3.12-7.18] | 3.54 [2.41-5.21] |
| *18C* | 8.14 [5.03-13.18] | 8.05 [5.51-11.76] | 10.10 [7.32-13.90] |
| *19A* | 12.73 [8.71-18.61] | 8.96 [6.43-12.49] | 10.10 [7.55-13.43] |
| *19F* | 8.47 [5.73-12.51] | 6.25 [4.54-8.62] | 6.00 [4.46-8.07] |
| *23F* | 13.24 [8.52-20.60] | 4.87 [2.96-8.00]*[a] | 3.07 [2.13-4.44]****[a] |

Concentrations are in ug/mL [95% CI]. [a]: compared to the young adults. *p<0.05, **p<0.01, ***p<0.001, ****p<0.0001

**Supplementary Table 3. Correlation between age and the 28 days post PCV13 vaccination pneumococcal serotype specific IgG concentrations.**

| *Pneumococcal serotype* | *rho* | *p-value* |
|---|---|---|
| *1* | -0.156 | 0.009* |
| *3* | -0.107 | 0.076 |
| *4* | -0.235 | <0.0001* |
| *5* | -0.183 | 0.002* |
| *6A* | -0.146 | 0.015* |
| *6B* | -0.208 | 0.001* |
| *7F* | -0.044 | 0.467 |
| *9V* | -0.074 | 0.219 |
| *14* | -0.050 | 0.413 |
| *18C* | 0.064 | 0.293 |
| *19A* | -0.025 | 0.680 |
| *19F* | -0.075 | 0.216 |
| *23F* | -0.251 | <0.0001* |

The Spearman rho is given. * indicates a significant correlation.

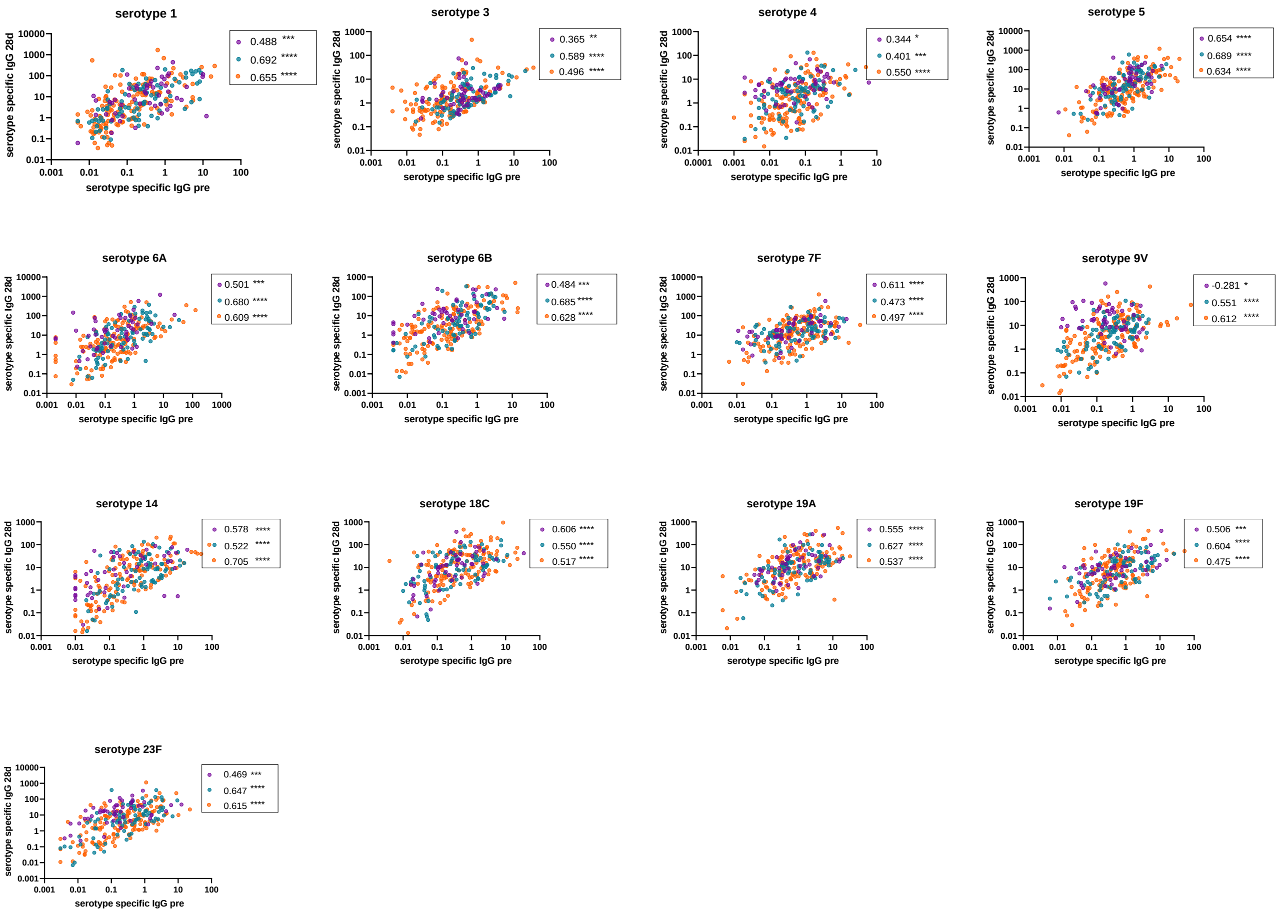


**Supplementary Figure 2. Analysis of the correlation between pre and post PCV13 vaccination serotype specific IgG concentrations.**

The correlation between the pre-vaccination and 28 days post PCV13 vaccination serotype specific IgG concentrations (ug/mL) (Log10 scale) separated for the 3 different age groups (young adults: purple, middle-aged adults: blue, older adults: orange). The Spearman's rho is indicated. *p<0.05, **p<0.01, ***p<0.001, ****p<0.0001.

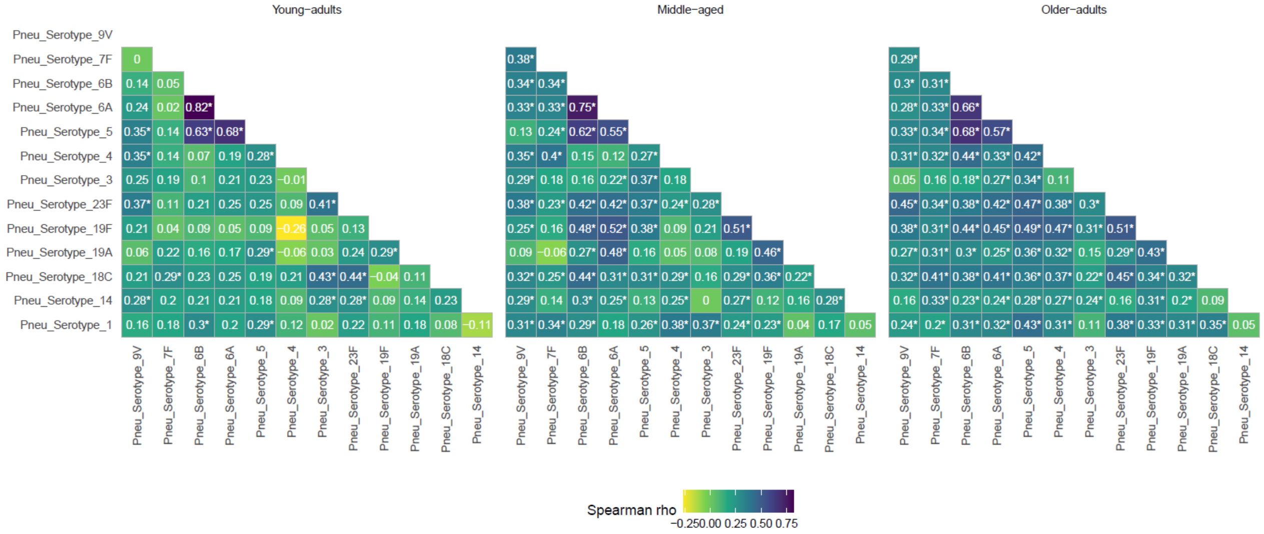


**Supplementary Figure 3. Partial correlation matrix between the pneumococcal serotype-specific IgG concentrations 28 days post-PCV13 vaccination split by age group.** The Spearman correlation coefficient is indicated and the * indicates a significant correlation.

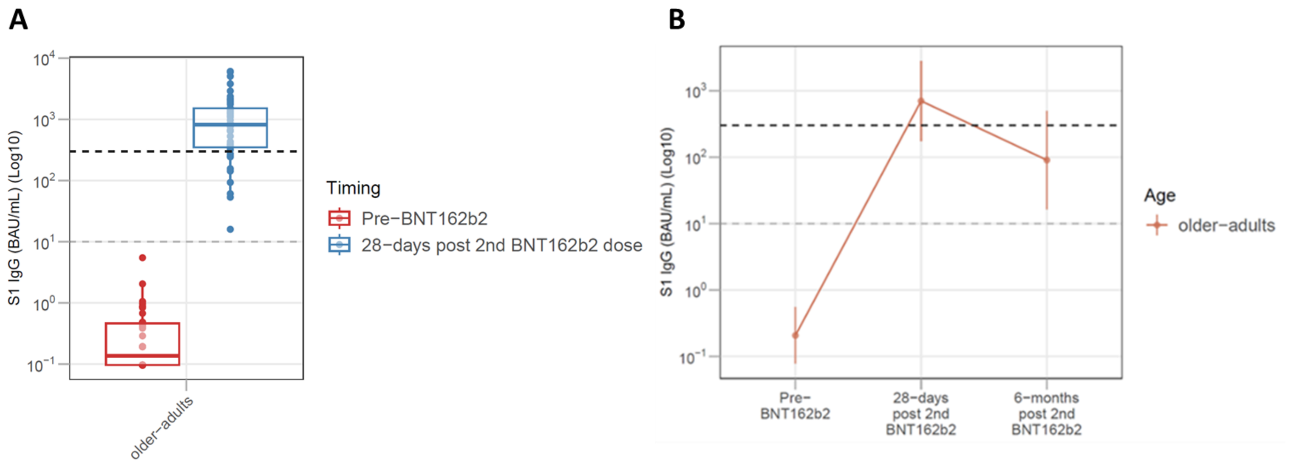


**Supplementary Figure 4. BNT162b2 induced S1 specific IgG responses in the older adults.**

**(A)** The pre- and 28- days post 2$^{nd}$ BNT162b2 vaccination S1-specific IgG concentrations (BAU/mL) (presented on a log10 scale) in the older adults (n=42) (mean age 84 (range 76-98)). The box plots indicate the median and interquartile range. **(B)** The longitudinal (pre-, 28 days, and 6 months post 2$^{nd}$ BNT162b2 vaccination) S1-specific IgG concentrations split in the older adults (n=39). The lines indicate the mean and standard error of the mean. No statistics are indicated in this graph. In both A and B, the grey dotted line (S1 specific IgG = 10 BAU/mL) indicates the cut-off for seropositivity. The black dotted line (S1 specific IgG = 300 BAU/mL) indicates the cut-off for an adequate response.

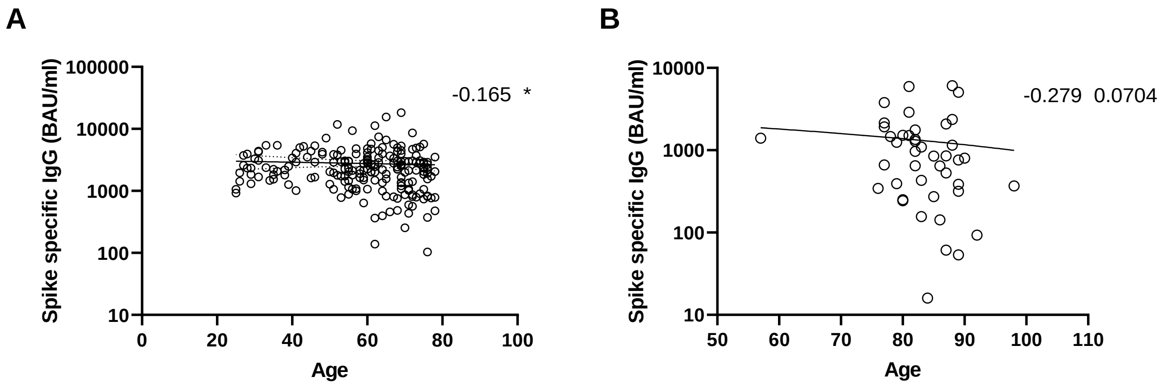


**Supplementary Figure 5. Analysis of SARS-COV2 vaccine responses in relation to age.**

The Spike specific S1 IgG concentrations (BAU/mL) following two vaccinations of mRNA-1273 **(A)** and BNT162b2 **(B)** in relation to age. The Spearman's rho is indicated. *p<0.05

**Supplementary Table 4. The division of the young, middle-aged and older adults over the different humoral response categories.**

| | **Young adults** (%) | **Middle-aged adults** (%) | **Older adults** (%) |
|---|---|---|---|
| | | **Dual vaccine analysis** | |
| | | *QIV* | |
| 1 | 14.6 | 26.5 | 30.1* |
| 2 | 29.2* | 28.9 | 23.5 |
| 3 | 29.2* | 21.7 | 20.6 |
| 4 | 27.1* | 22.9 | 25.7 |
| | | *PCV13* | |
| 1 | 8.3 | 26.5 | 30.9* |
| 2 | 18.8 | 20.5 | 27.9* |
| 3 | 47.9* | 30.1 | 11.8 |
| 4 | 25 | 22.9 | 29.4* |
| | | *Dual score* | |
| 1 | 0 | 6 | 11* |
| 2 | 27.1 | 32.5* | 29.4 |
| 3 | 43.8* | 43.4 | 41.9 |
| 4 | 29.2* | 18.1 | 17.6 |
| | | **Triple vaccine analysis** | |
| | | *QIV* | |
| 1 | 12.2 | 22.9 | 34.2* |
| 2 | 31.7* | 30.0 | 21.5 |
| 3 | 34.1 | 37.1* | 26.6 |
| 4 | 22.0* | 10.0 | 17.7 |
| | | *PCV13* | |
| 1 | 12.2 | 27.1 | 31.6* |
| 2 | 29.3 | 22.9 | 30.4* |
| 3 | 24.4 | 25.7* | 10.1 |
| 4 | 34.1* | 24.3 | 27.8 |
| | | *mRNA-1273* | |
| 1 | 12.2 | 21.4 | 35.4* |
| 2 | 31.7* | 27.1 | 19.0 |
| 3 | 22.0 | 28.6* | 22.8 |
| 4 | 34.1* | 22.9 | 22.8 |
| | | *Triple score* | |
| 1 | 2.4 | 8.6 | 12.7* |
| 2 | 31.7 | 42.9 | 49.4* |
| 3 | 53.7* | 42.9 | 31.6 |
| 4 | 12.2* | 5.7 | 6.3 |

The division of participants of every age group over the 4 categories is calculated per vaccine or response score. The age group with the highest proportion in a response category is indicate with a *.
